# Trends and hotspots in research on osteoporosis and nutrition from 2004 to 2024: a bibliometric analysis

**DOI:** 10.1101/2024.04.14.24305794

**Authors:** Min Li, Binyang Yu, Haiyan Yang, Haiyan He, Ning Li, Aili Lv, Xiaoling Zhou, Rui Gao

**Author notes:** (R.G.). Min Li and Binyang Yu contributed equally to this work.

## Abstract

**Objectives:** This study aimed to perform a bibliometric analysis utilizing the Web of Science database on osteoporosis and nutrition-related research published from 2004 to 2024.

**Background:** In recent years, the intricate association between nutrition and osteoporosis has garnered increasing attention. However, there is currently no published bibliometric research on this topic.

**Methods:** The data were extracted from the Web of Science Core Collection (WOSCC) from inception to 2024-01-31 and analyzed using bibliometric methods and CiteSpace, incorporating variables such as annual publication volume, author patterns, institutional affiliations, country/region contributions, journal publications, highly cited literature, and keyword clustering.

**Results:** A total of 2138 articles were assessed, revealing a consistent upward trend in published works in this domain, with the majority originating from the United States. Seoul National University was identified as the most prolific institution. Among the authors, Geng, Bin were the most prolific, while Kanis J.A. garnered the highest citation count. Research hotspots included bone density, postmenopausal women, vitamin D, hip fractures, etc. Research subjects included physical activity, sarcopenia, calcium intake, machine learning, etc.

**Conclusions:** Our comprehensive analysis offers overviews of research trends and hotspots in the field of osteoporosis and nutrition over the past two decades, highlighting some shortcomings and hopefully providing valuable insights and guidance for future researchers and decision-makers.

## Introduction

Osteoporosis is defined as reduced bone mineral density (BMD) and increased bone fragility[1]. It ranks among the most common ailments affecting middle-aged and elderly individuals according to the World Health Organization(WHO)[2]. Globally, its prevalence is 18.3%, with a higher incidence in women than in men (23.1% and 11.7%, respectively)[3,4]. Osteoporosis manifests primarily as the loss of bone mass, degradation of bone microstructure, increased bone fragility, and a heightened risk of fractures[1]. In severe cases, fractures may occur, leading to elevated disability rates, depression, diminished quality of life, and even mortality[5]. Previous studies have shown that more than one-third of middle-aged and elderly women (compared to one in five men) in the world suffer from fractures due to the effects of osteoporosis[6,7]. By 2050, the number of hip fractures is projected to surpass 21 million[8]. This alarming trend toward osteoporosis is exacerbated by the aging population, as evidenced by the increasing incidence of clinical cases in middle-aged men. With the increase in the age of Americans according to census population projections, the incidence of osteoporosis is expected to increase by 32% to 17.2 million from 2010 to 2030[9]. Moreover, osteoporosis imposes a substantial economic burden worldwide. In the European Union alone, the estimated cost of preventing and managing osteoporosis amounts to €37 billion, a figure set to increase by 25% by 2025[10]. Therefore, osteoporosis has become a major public health problem worldwide[11].

Bone, as a living tissue, relies on a full spectrum of essential nutrients for growth and maintenance. It consists predominantly of protein, the principal component of connective tissue, which constitutes 50% of bone volume and 20% of bone weight. Good nutrition plays a pivotal role to maintain optimal bone health. Consistent small benefits gained daily over the course of several decades can significantly impact one’s fracture risk[12]. Nutrition, in particular, has emerged as a key determinant in mitigating bone loss and the risk of fractures. Several studies strongly recommend a diet rich in calcium, protein, vitamin C, and vitamin D as essential for preventing osteoporosis[13–15]. Previous research has substantiated the correlation between micronutrients and the prevention of osteoporosis, emphasizing the significance of dietary factors[16]. Conversely, poor nutrition has been identified as a contributing factor to osteoporosis and fractures[17].

Osteoporosis has emerged as a significant clinical challenge confronting the global human population. Understanding the intricate relationship between osteoporosis and nutrition is imperative for the development of effective preventive and therapeutic interventions. Despite the universal attention bestowed upon the relationship between osteoporosis and nutrition topic, despite the existence of approximately 600 reviews and over 3000 original research studies in the field. Bibliometrics, as a quantitative method, employs mathematical and statistical approaches to analyze scientific publications. By doing so, it provides a comprehensive overview of contributions distribution, identifies hotspots, and reveals future trends within a specific area of study. CiteSpace, in particular, facilitates the conceptualization of knowledge domains by generating and visualizing co-occurrence network maps of contributors and keywords as well as co-citation networks of cited authors.

Thus, the objective of this study was to conduct a bibliometric analysis of osteoporosis and nutrition studies published from 2004 to 2024 utilizing data obtained from the Web of Science database. The comprehensive analysis might form the basis for evidence-based strategies aimed at promoting bone health and alleviating the substantial global burden caused by osteoporosis. Consequently, it is hoped that this study will provide crucial insights and guidance for future researchers and decision makers, facilitating advancements in the field.

## Materials and Methods

### Data source and search strategy

The research process is illustrated in Fig 1. We collected the data from the Web of Science Core Collection database. The timespan covered from 2004.01.01 to 2024.01.31. The “topic” field was used to search for articles related to a specific research field. TS=(“Nutrition” or “Nutrients” or “Nutritional Supplements” or “Dietary Nutrients” or “Dietary Supplementations” or “Macronutrients” or “Micronutrients”) and (“Osteoporosis, Postmenopausal” or “Osteoporosis” or “Posttraumatic Osteoporosis” or “Senile Osteoporosis” or “Age Related Bone Loss” or “Age Related Osteoporosis”).

**Fig 1.**
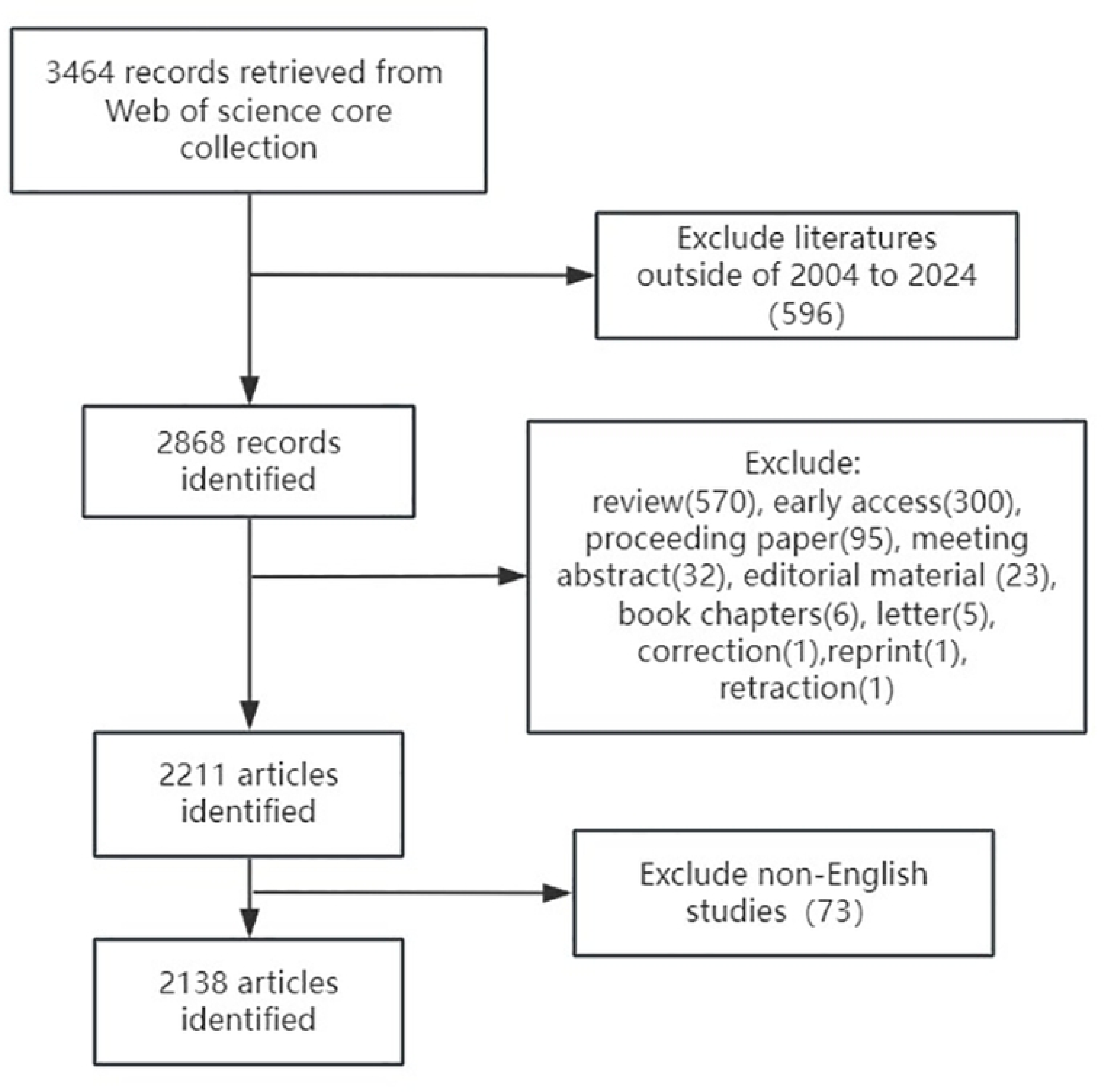
Flow chart of the inclusion criteria.

### Data analysis and visualization

CiteSpace, developed by Chaomei Chen, is a Java application designed to visualize bibliographic databases effectively. In this study, we utilized CiteSpace 6.1. R2 software to map scientific knowledge, employing co-occurrence and clustering analyses on high-frequency terms, countries, etc. The software was configured with the following specific settings: a time slicing range from 2004 to 2024, with one year per slice; selection of author, institution, country, keyword, reference, cited author, and cited journal as term sources; and pathfinder and pruning sliced networks for network pruning. A total of 2138 papers were included in the study. Trend maps representing publication volume, authorship, institutional cooperation, geographical distribution, and keyword mapping were generated to highlight key nodes and research hotspots, effectively scientific research and can be selectively analyzed based on specific problem domains[18]. Furthermore, during the mapping process, CiteSpace provides module values (Q-values) and average profile values (S-values) to ensure the effectiveness of the mapping. A Q value greater than 0.3 indicates a significant network structure, while an S value greater than 0.5 suggests reasonable clustering, and an S value greater than 0.7 signifies plausible clustering[18]. CiteSpace facilitates the presentation of domain-specific research trends through quantitative analysis and visual representation, thereby revealing the development, hotspots, and frontiers of scientific research in the field.

## Results

### Analysis of annual publications

In this study, a comprehensive analysis was conducted on 2138 articles spanning the last two decades in the field of osteoporosis and nutrition. Fig 2 provides a clear visualization of the upward trend in publication numbers. It can be observed that the number of publications exhibited a gradual increase over time, starting from 46 in 2004 and reaching a peak of 205 in 2023, indicating the growing interest of researchers in this relevant topic. Notably, the rate of increase intensified after 2018, further emphasizing the increasing attention dedicated to this subject matter.

**Fig 2.**
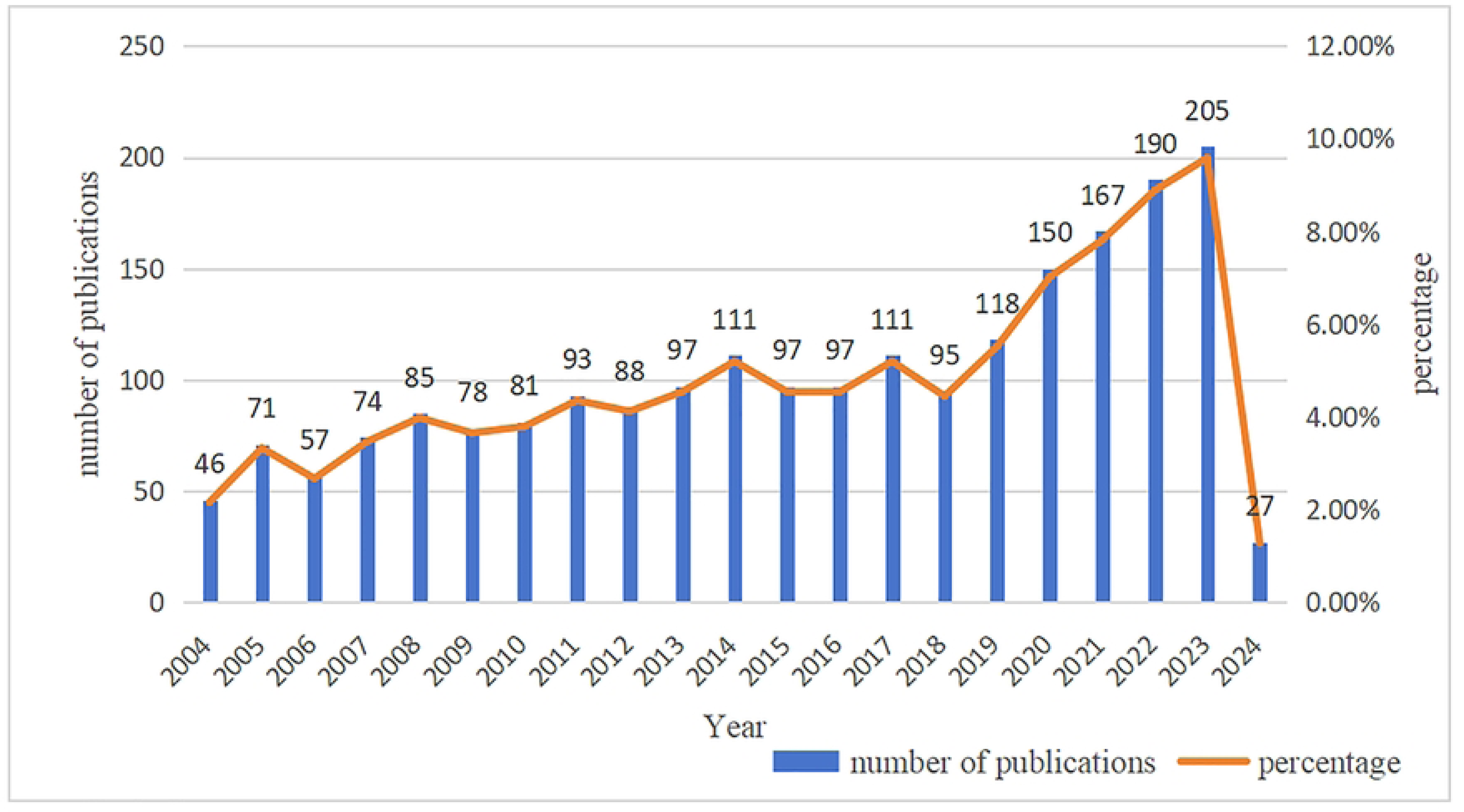
Trend of annual publications from 2004 to 2024.

### Analysis of countries and institutions

As indicated in Tables 1 and 2, in the field of osteoporosis and nutrition, the United States was the most active participant and was responsible for the highest number of published papers, encompassing 564 papers (21%) of the total. China occupies the second position with 317 papers, accounting for 11.8% of the cumulative count. South Korea ranks third with 285 articles, followed by England, Canada, Japan, Italy, Spain, Australia, and Germany. Notably, universities emerge as the predominant institutions generating these publications, with Seoul National University (SNU), Harvard University, and the University of California System reigning supreme in contribution. The respective publication counts for these institutions are 56, 49, and 48 in descending order. Several other influential research institutions include the United States Department of Agriculture (USDA), Catholic University of Korea, Harvard Medical School, and Yonsei University. Generally, these institutions predominantly hail from the United States, South Korea, and Canada. Fig 3 visually represents the international collaboration in the realm of osteoporosis and nutrition, showcasing 86 nodes connected by 437 lines with a network density of 0.1196. This finding signifies that researchers from numerous countries are actively engaging in studies pertaining to osteoporosis

**Table 1.** Top 10 countries/regions by the number of publications.

| Rank | Country/Region | Publicatons | Percentage(%) |
| --- | --- | --- | --- |
| 1 | United States | 564 | 21.0% |
| 2 | China | 317 | 11.8% |
| 3 | South Korea | 285 | 10.6% |
| 4 | England | 148 | 5.5% |
| 5 | Canada | 114 | 4.2% |
| 6 | Japan | 86 | 3.2% |
| 7 | Italy | 83 | 3.1% |
| 8 | Spain | 82 | 3.1% |
| 9 | Australia | 79 | 2.9% |
| 10 | Germany | 62 | 2.3% |

**Table 2.** Top 10 institutions by the number of publications.

| Rank | Institutions | Country/Region | Publicatons |
| --- | --- | --- | --- |
| 1 | Seoul National University (SNU) | South Korea | 56 |
| 2 | Harvard University | United States | 49 |
| 3 | University of California System | United States | 48 |
| 4 | United States Department of Agriculture (USDA) | United States | 35 |
| 4 | Catholic University of Korea | South Korea | 35 |
| 5 | Harvard Medical School | United States | 32 |
| 6 | Yonsei University | South Korea | 28 |
| 6 | Centers for Disease Control & Prevention - USA | United States | 28 |
| 6 | Tufts University | United States | 28 |
| 7 | University of Toronto | Canada | 26 |

**Fig 3.**
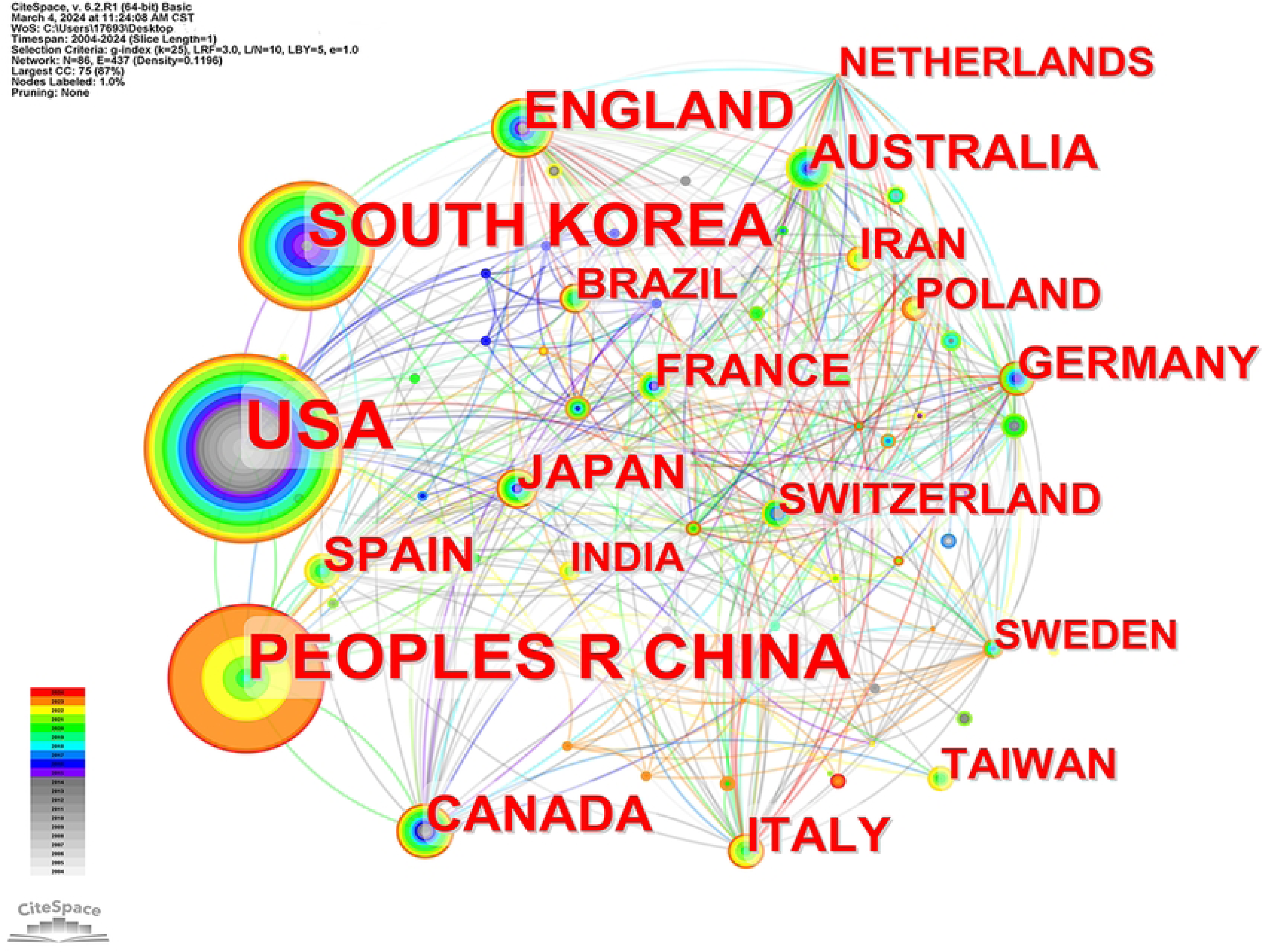
Country/region collaboration chart.

### Analysis of cited journals

Table 3 presents the top 10 journals in terms of citation volume, with the majority hailing from the United States, except for Osteoporosis International and The Lancet, which originated from Germany and England, respectively. The three leading journals in this ranking are Osteoporosis International, Journal of Bone and Mineral Research, and The American Journal of Clinical Nutrition. Their respective citation counts are 1449, 1366, and 1115, with corresponding impact factors of 4.0, 6.2, and 7.1, respectively. Notably, The Lancet and The New England Journal of Medicine had the highest impact factors at 168.9 and 158.5, respectively. Consequently, it can be inferred that this field boasts a selection of high-quality articles. Fig 4 illustrates the collaboration among cited journals and reveals 826 nodes connected by 4310 lines, generating a network density of 0.0126. In this visualization, the journals ranked at the top exhibit larger nodes, indicating their prominence in terms of collaboration and citation impact.

**Table 3.** Top 10 journals by citations.

| Rank | Cited journals | Citations | Country | Category zone | IF(2023) |
| --- | --- | --- | --- | --- | --- |
| 1 | Osteoporosis International | 1449 | Germany | Q2 | 4.0 |
| 2 | Journal of Bone and Mineral Research | 1366 | United States | Q1 | 6.2 |
| 3 | The Amercian Journal of Clinical Nutrition | 1115 | United States | Q1 | 7.1 |
| 4 | Bone | 1032 | United States | Q2 | 4.398 |
| 5 | The Journal of Clinical Enocrinology and Metabolism | 983 | United States | Q1 | 5.8 |
| 6 | Calcified Tissue International | 739 | United States | Q3 | 4 |
| 7 | The New England Journal of Medicine | 682 | United States | Q1 | 158.5 |
| 8 | The Lancet | 681 | England | Q1 | 168.9 |
| 9 | Journal of Nutrition | 663 | United States | Q2 | 4.687 |
| 10 | JAMA-Journal of the American Medical Directors Association | 635 | United States | Q1 | 7.6 |

**Fig 4.**
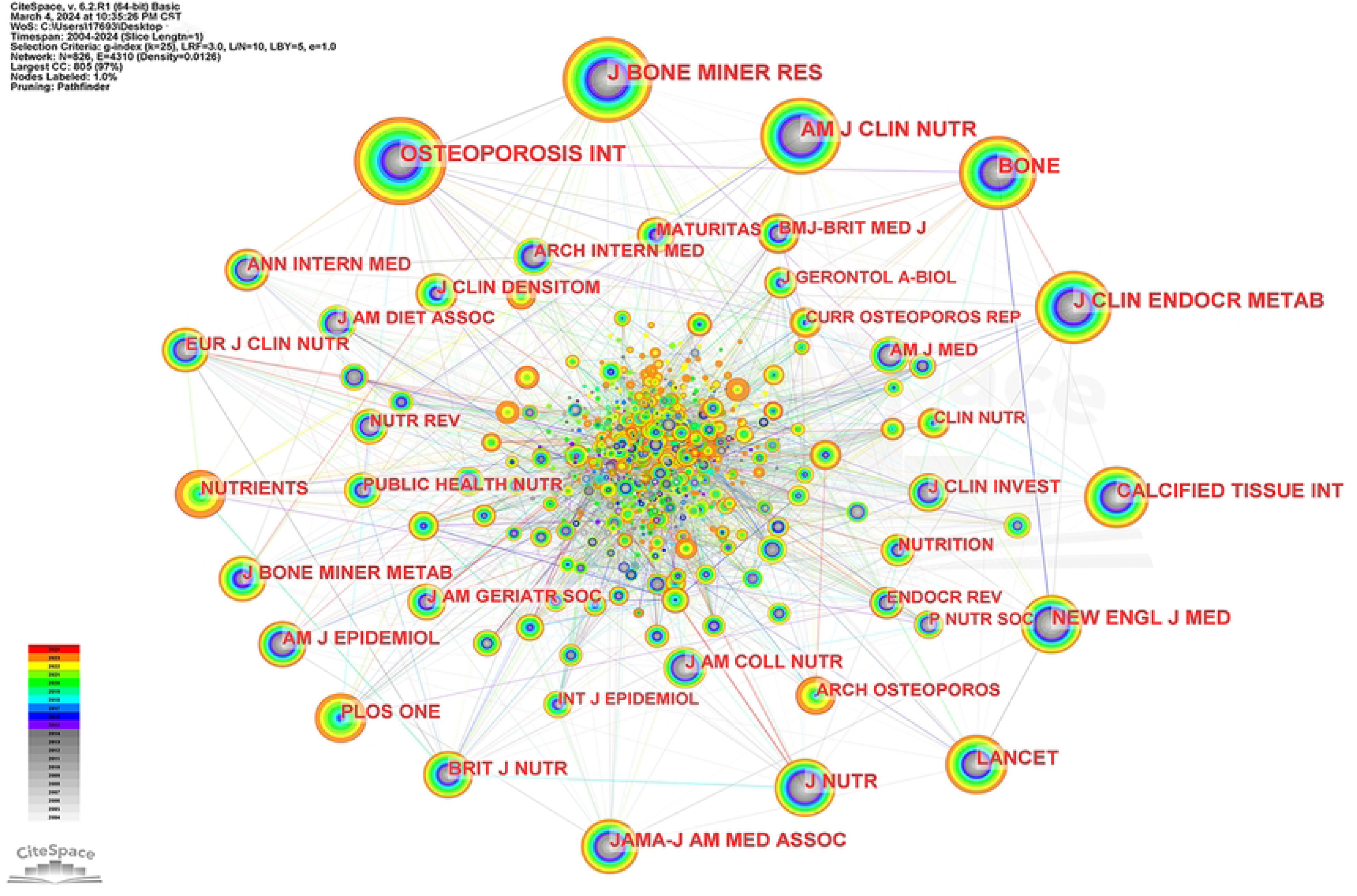
Collaboration chart of cited journals.

### Analysis of authors and co-cited authors

The data presented in Tables 4 and 5 reveal that the majority of the top 10 authors in terms of publication volume originate from China, followed by the United States and South Korea. The number of authors with the highest number of publications was 12, 11, and 10, respectively. Notably, three scholars, namely, Liu Mingjiang, Zhang Ya, and Xie Ruijie, represent the same institution and exhibit close collaboration among themselves. By analyzing co-cited frequencies, we found that the three authors with the highest occurrences are Kanis J.A., Heaney R.P., and Looker A.C. These scholars hail from England and the United States, and their co-cited frequencies are 436, 264, and 242, respectively. It is worth noting that Looker A.C., in addition to contributing significantly to the publication volume, also has a high frequency of co-citation, implying substantial influence in this field. The number and size of the nodes in the author and co-cited author collaboration network map demonstrate the frequency of co-occurrence, while the connected lines indicate the strength of the collaborative network among authors. In Fig 5, the author collaboration network comprises 318 nodes and 528 connected lines, resulting in a network density of 0.0105. Similarly, Fig 6 shows the co-cited author collaboration network, composed of 834 nodes and 2132 connected lines, with a network density of 0.0061.

**Table 4.** Top 10 active authors in the research.

| Rank | Author | Publications | Country | Affiliation |
| --- | --- | --- | --- | --- |
| 1 | Geng, Bin | 12 | China | Department of Orthopedics, Lanzhou University Second Hospital |
| 2 | Looker, A C | 11 | United States | Division of Health and Nutrition Examination Surveys, National Center for Health Statistics, Centers for Disease Control and Prevention |
| 3 | Rhee, Yumie | 10 | Korea | Department of Internal Medicine, Yonsei University College of Medicine |
| 3 | Xia, Yayi | 10 | China | Department of Orthopedics, Lanzhou University Second Hospital |
| 3 | Tang, Yuchen | 10 | China | Chongqing Medical University |
| 4 | Liu, Mingjiang | 9 | China | Nanhua Hospital, Hengyang Medical School, University of South China |
| 4 | Zhu, Zhongxin | 9 | China | Hospital of Wenzhou Medical University |
| 5 | Zhang, Ya | 8 | China | Nanhua Hospital, Hengyang Medical School, University of South China |
| 5 | Weaver, Connie M | 8 | United States | Department of Nutrition and Food Studies, George Mason University |
| 5 | Xie, Ruijie | 8 | China | Nanhua Hospital, Hengyang Medical School, University of South China |

**Table 5.** Top 10 most co-cited authors.

| Rank | Co-cited author | Co-cited frequency | Country | Affiliation |
| --- | --- | --- | --- | --- |
| 1 | Kanis J.A. | 436 | England | Centre for Metabolic Bone Diseases, University of Sheffield Medical School |
| 2 | Heaney R.P. | 264 | United States | Department of Medicine, Creighton University |
| 3 | Looker A.C. | 242 | United States | Division of Health and Nutrition Examination Surveys, National Center for Health Statistics, Centers for Disease Control and Prevention |
| 4 | Holick M.F | 193 | United States | Boston University School of Medicine |
| 5 | Reid I.R. | 154 | New Zealand | University Auckland |
| 5 | Cummings S.R. | 154 | United States | California Pacific Medical Center Research Institute San Francisco Coordination Center |
| 6 | Bischoff-Ferrari H.A. | 150 | Switzerland | Department of Aging Medicine and Aging Research, University Hospital of Zurich, Switzerland |
| 7 | Rizzoli R. | 140 | Switzerland | Bern University Hospital, University of Bern |
| 8 | Johnell O. | 136 | Sweden | Karolinska Institute, Department of Epidemiology and Biostatistics Medicine |
| 9 | Tucker K.L. | 132 | England | Nuffield Department of Primary Health Care Sciences, University of Oxford |

**Table 6.**
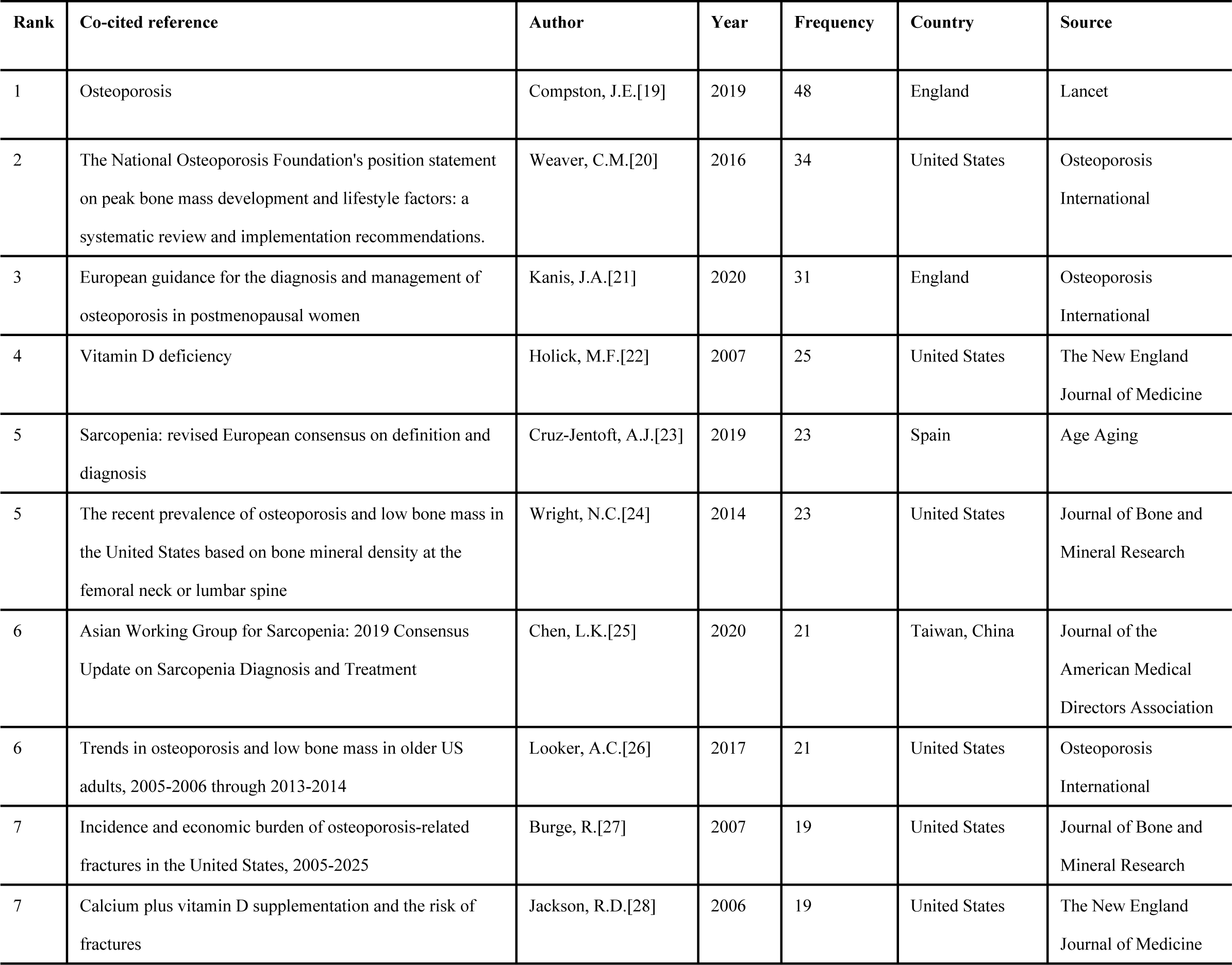
Top 10 co-cited references in the research.

**Table 7.** Co-occurrence frequency of the top 10 keywords.

| Rank | Keywords | Number of occurrences | Centrality |
| --- | --- | --- | --- |
| 1 | osteoporosis | 788 | 0 |
| 2 | bone mineral density | 731 | 0 |
| 3 | women | 387 | 0.01 |
| 4 | postmenopausal women | 335 | 0.02 |
| 5 | risk | 318 | 0.02 |
| 6 | vitamin D | 290 | 0.01 |
| 7 | mineral density | 284 | 0.01 |
| 8 | health | 259 | 0.02 |
| 9 | mass | 236 | 0.01 |
| 10 | hip fracture | 214 | 0.03 |

**Fig 5.**
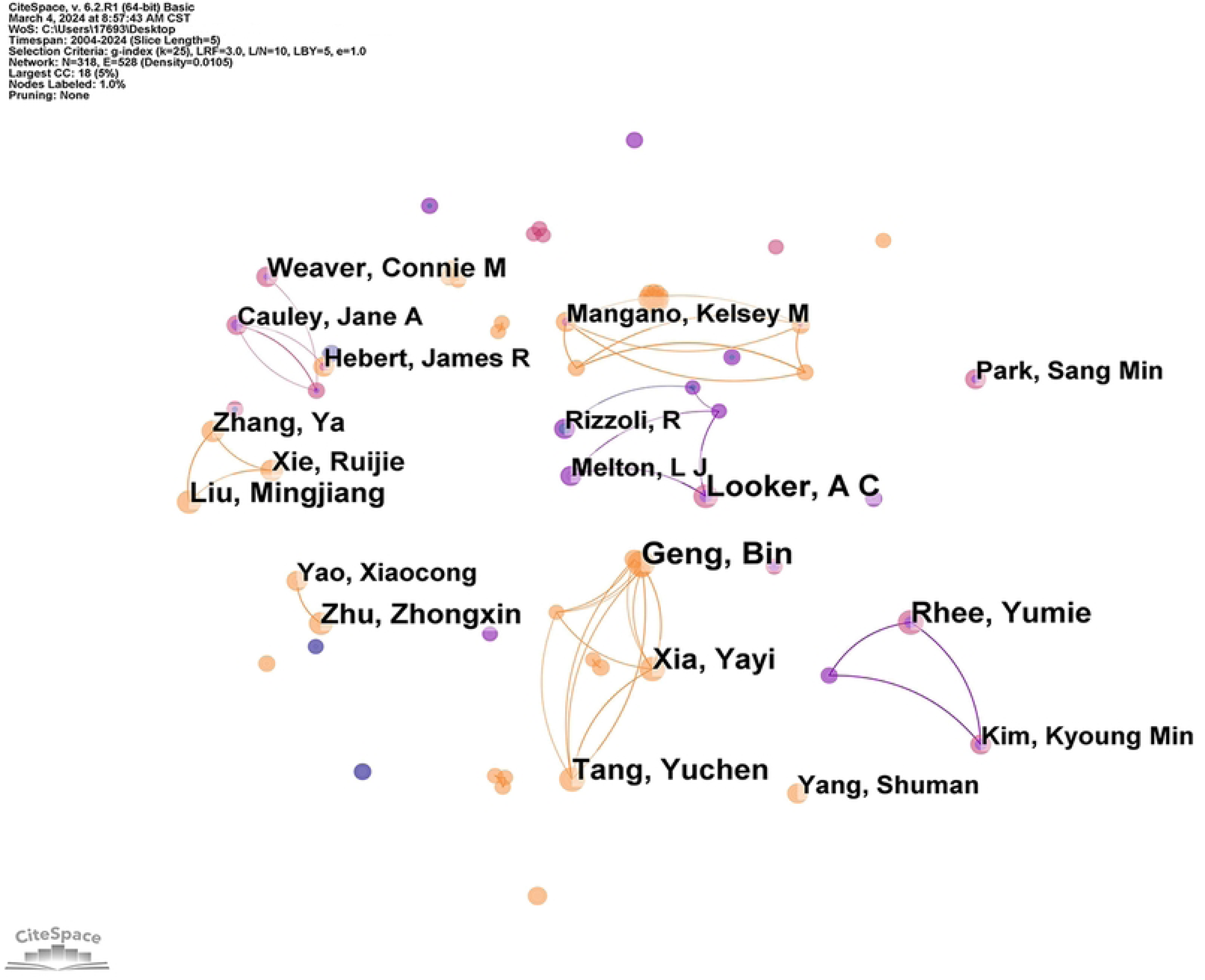
Author collaboration chart.

**Fig 6.**
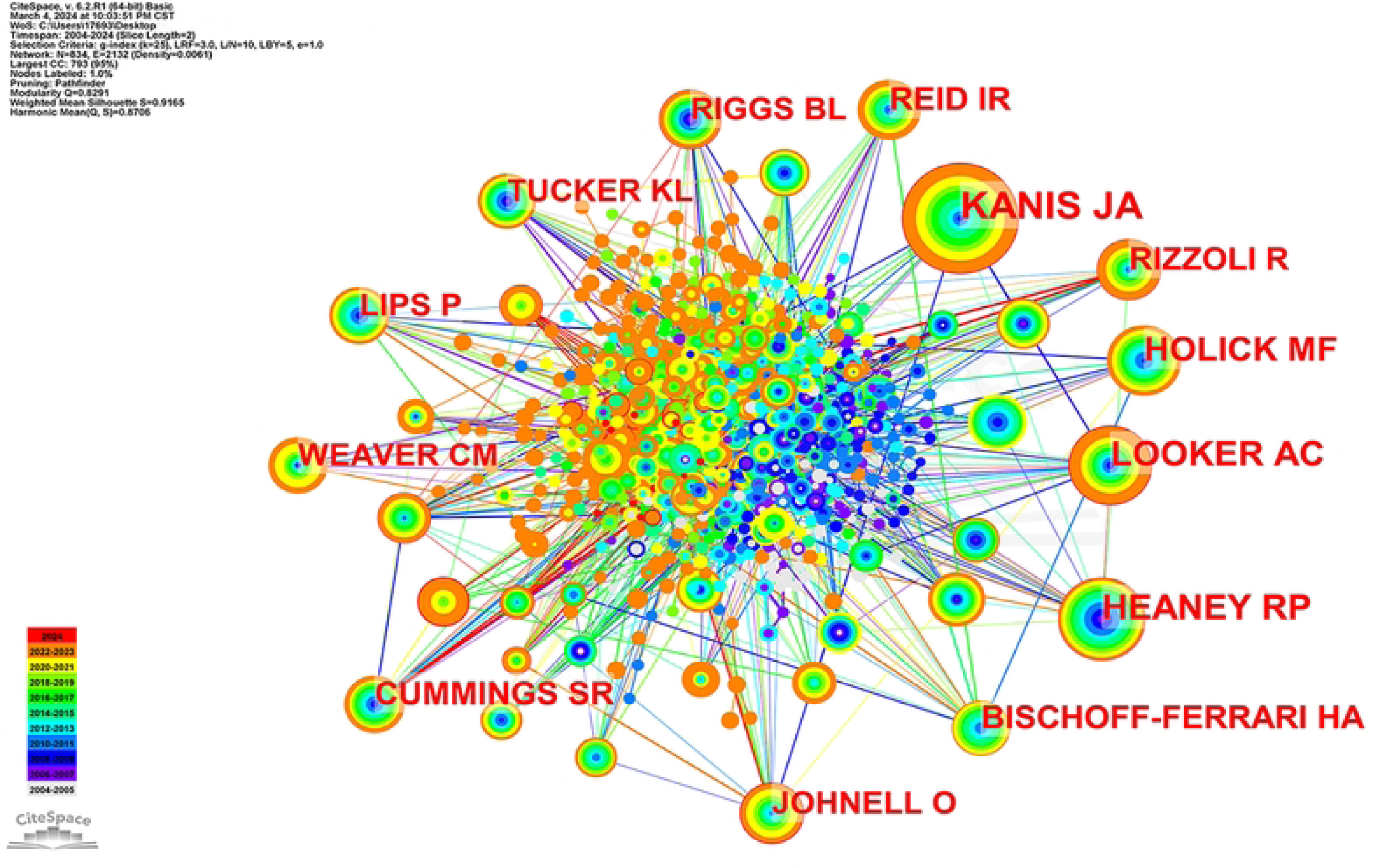
Collaboration chart of co-cited authors.

### Analysis of co-cited references

Table 6 provides a comprehensive summary of the main information pertaining to the top 10 most co-cited references. The most frequently co-cited reference is the article “Osteoporosis”. Authored by Compston, J.E. from the Department of Medicine at Cambridge Biomedical Campus in England, this article was published in Lancet in 2019 and has garnered 48 citations. It is worth mentioning that a majority of the authors of these highly cited articles are from the United States, with the remaining contributors hailing from England, Spain, and Taiwan, China. Among the top 10 cited articles, the second and third positions are occupied by articles from “Osteoporosis International”, while the fourth and last positions belong to articles from “The New England Journal of Medicine”. Additionally, the sixth and ninth positions are held by articles from the “Journal of Bone and Mineral Research”. Fig 7 presents the co-cited reference network, which consists of references with higher centrality and citation counts. This network aids in identifying the pivotal knowledge base within the field, offering convenience in

**Fig 7.**
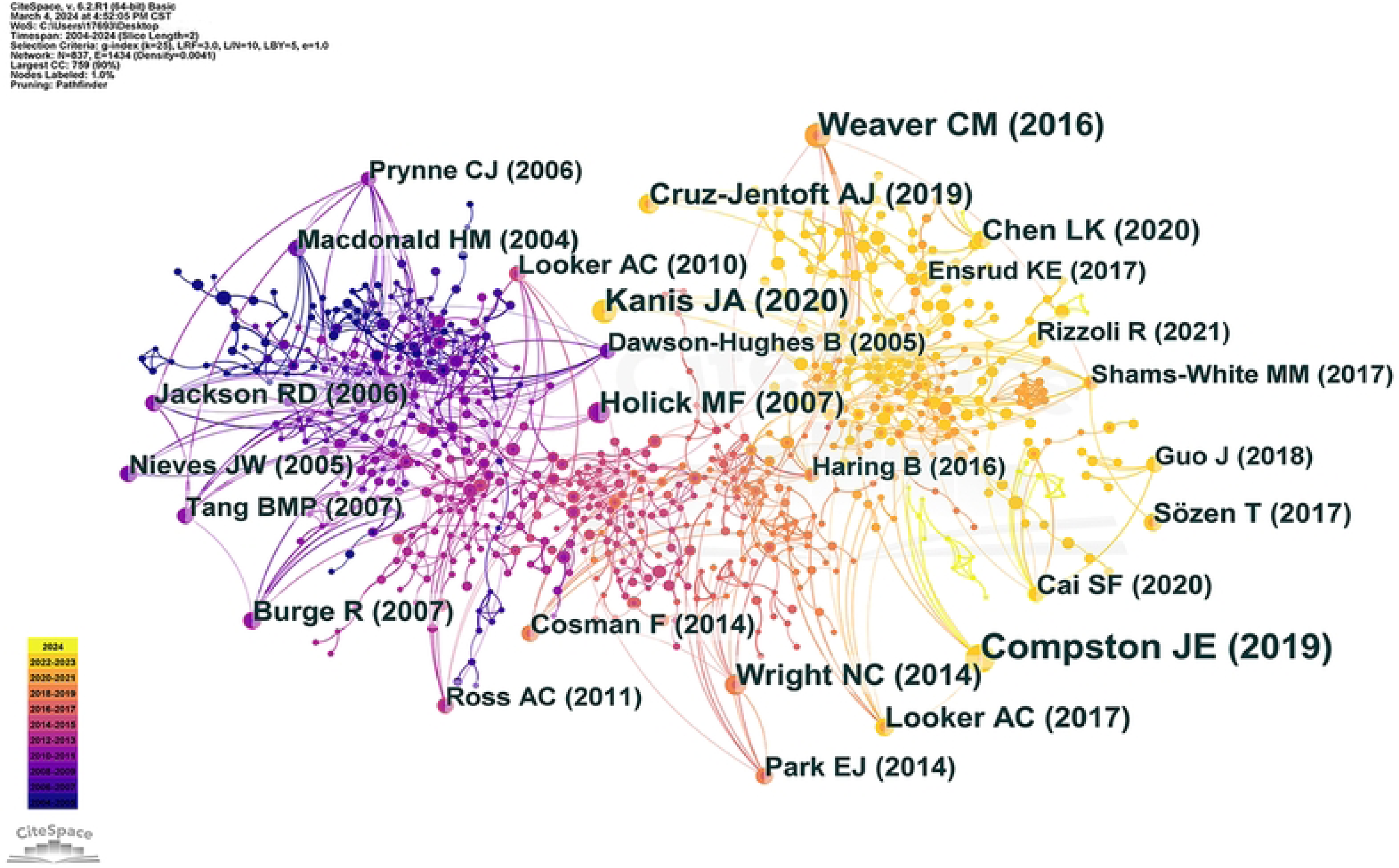
The collaboration chart of co-cited references.

### Analysis of keywords

#### Keyword co-occurrence analysis

To identify the hotspots and frontiers of retracted publications spanning from 2004 to 2024, a comprehensive analysis of keyword co-occurrence is imperative. Consequently, CiteSpace was employed to construct a keyword knowledge co-occurrence map, as shown in Fig 8. The time slice was set to 1 year, resulting in 624 nodes connected by 3718 links, with a network density of 0.0191. The size of each node corresponds to the significance of the respective keyword. To provide a more insightful understanding of these keywords, we present the ten most frequently occurring terms, along with their “postmenopausal women”, “risk”, “vitamin D”, “mineral density”, “health”, “mass”, and “hip fracture”. These keywords offer valuable insights into the prevailing themes and concerns within the research field.

**Fig 8.**
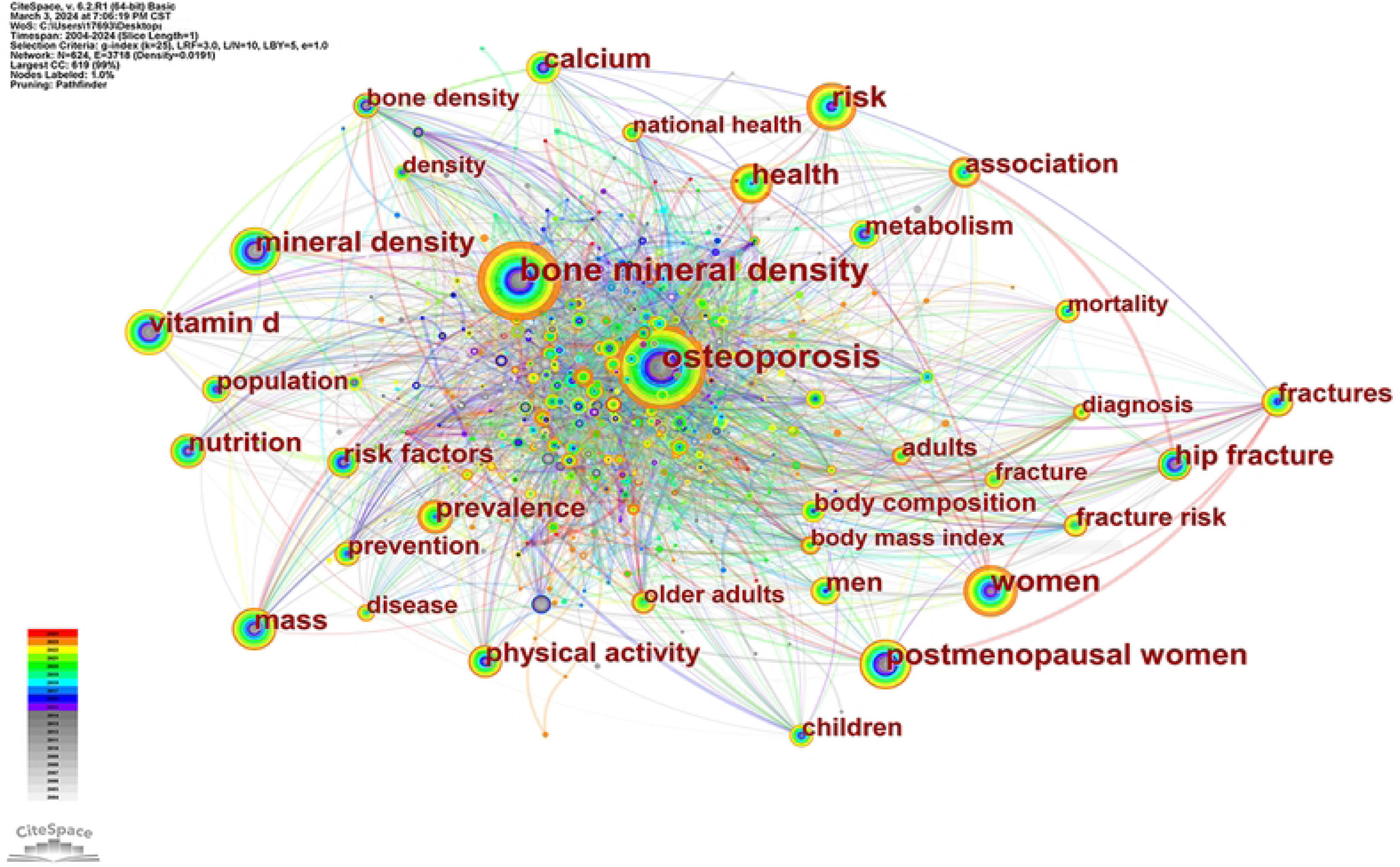
Keyword co-occurrence knowledge map.

#### Keyword clustering and citation bursts analysis

To gain a comprehensive understanding of the current research hotspots in the field, CiteSpace visualization software was used to generate a keyword clustering map. The scientific nature of the clustering map can be primarily deduced from the module value (Q value) and the average profile value (S value). In our study, a Q value of 0.3351, indicative of a favorable clustering effect, and an S value of 0.6812, indicating high homogeneity among the clusters and reasonable clustering outcomes, were obtained. Moreover, it is noteworthy that certain mapping clusters exhibit overlapping, suggesting their close correlation. As depicted in Fig 9, the major clusters encompass various areas, including #0 physical activity, #1 sarcopenia, #2 machine learning, #3 vitamin D, #4 metabolism, #5 body mass index, #6 nutrition examination survey, #7 trabecular bone, and #8 quality of life. Additionally, an analysis of keywords in CiteSpace was conducted to identify terms with significant citation bursts, which serve as indicators of emerging frontiers in a given period. Fig 10 presents the top 20 keywords characterized by the highest citation bursts that persist until 2024, offering a foundational understanding of recent frontiers and potential future directions. The top 10 keywords were “elderly women”, “calcium supplementation”, “3rd national health”, “blood pressure”, “hypovitaminnosis d”, “calcium intake”, “premenopausal”, “d insufficiency”, “biochemical markers”, and “dietary calcium”.

**Fig 9.**
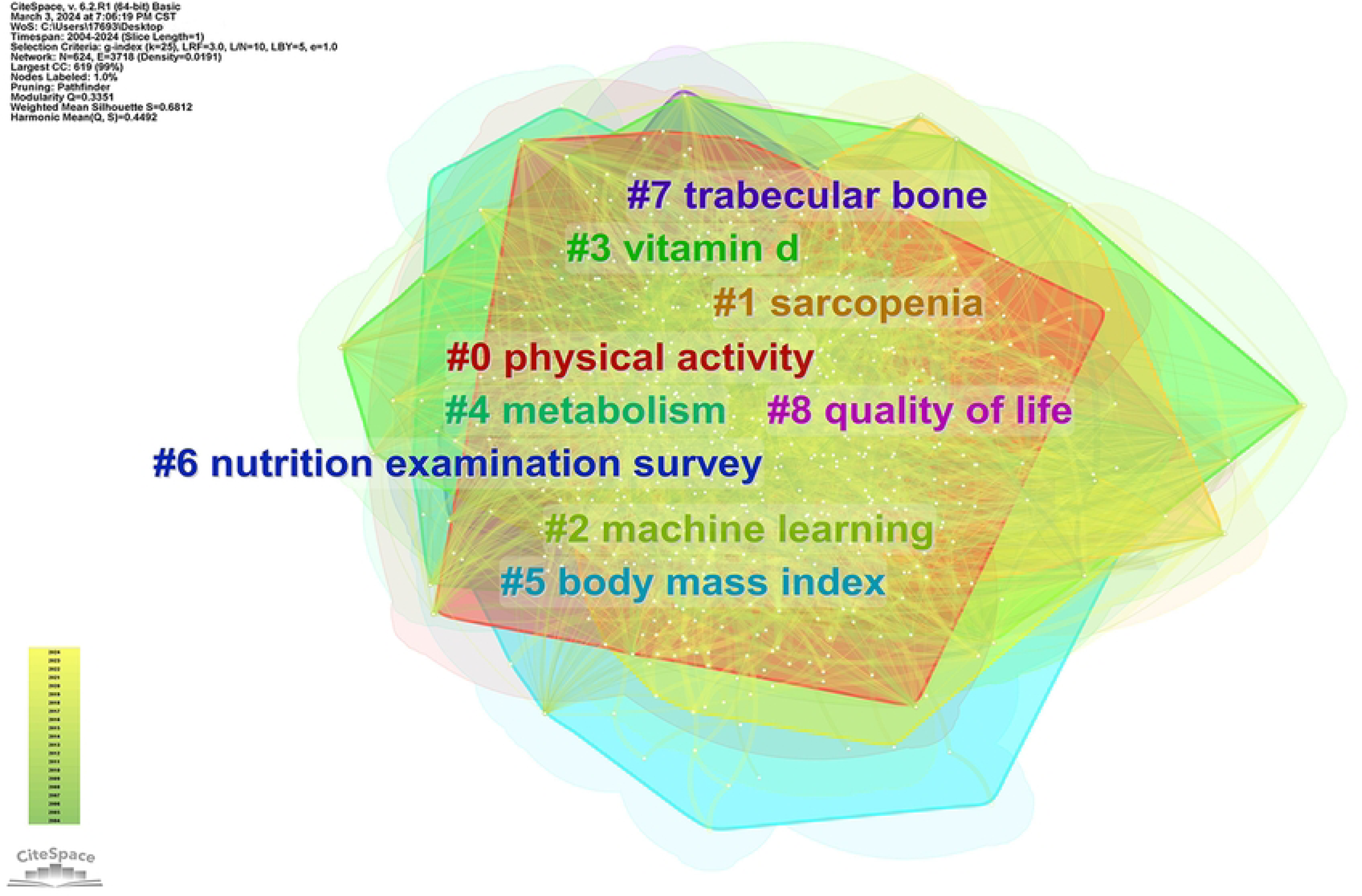
Keyword clustering map.

**Fig 10.**
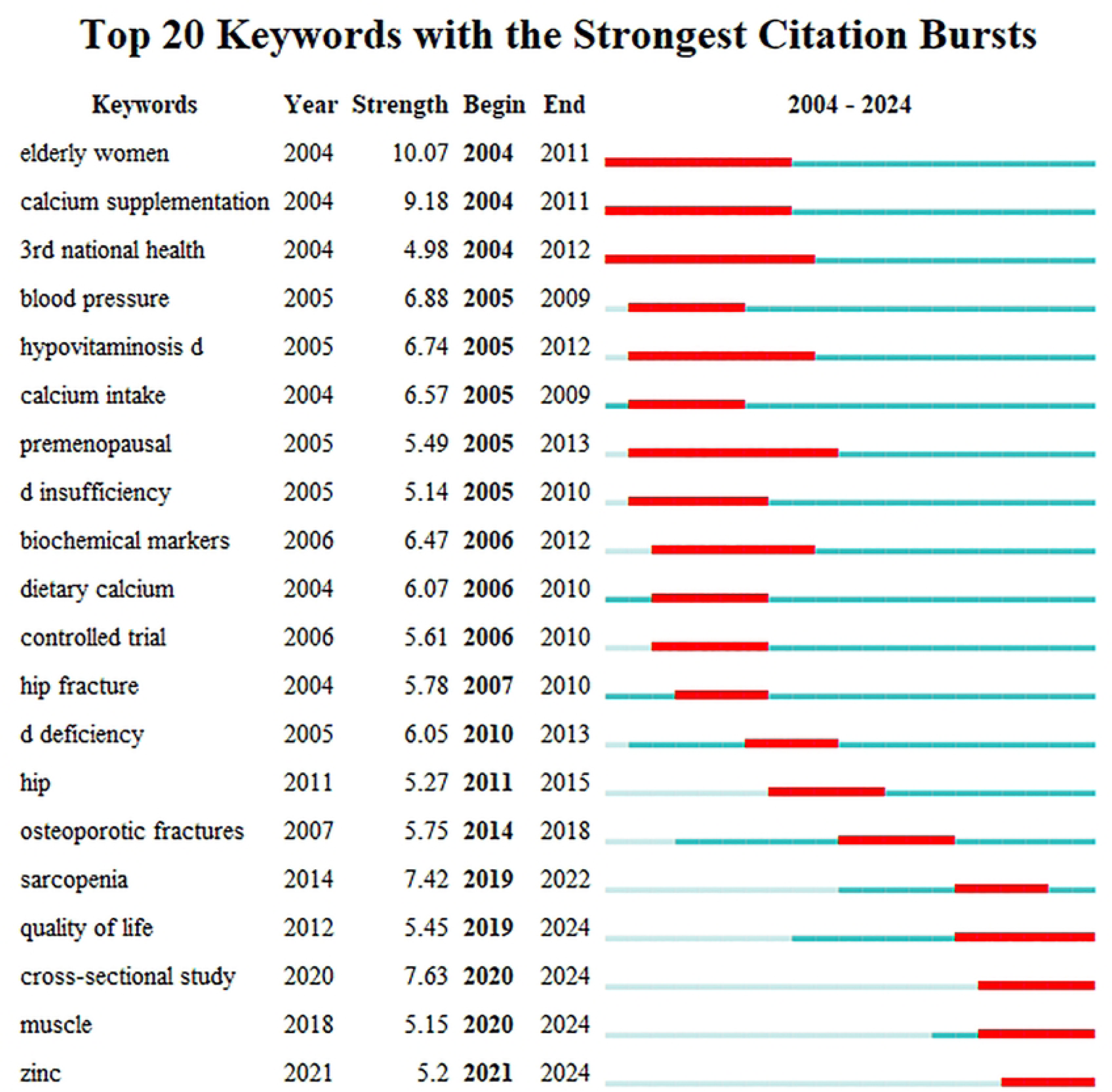
Keywords citation bursts.

## Discussion

### Principal Findings

This research employed CiteSpace software to conduct bibliometric analysis of published articles pertaining to osteoporosis and nutrition studies from 2004 to 2024. The authors, institutions, journals, countries, references, and keywords associated with osteoporosis and nutrition were examined to determine the current status and development trends. The analysis revealed a consistent and significant increase in the number of publications over time, indicating that sustained attention has been devoted to researching osteoporosis and nutrition in recent years. Several factors contribute to this upsurge in interest. First, the growing focus on health and nutrition, coupled with advancements in research methods and tools, has prompted researchers to extensively and deeply explore the intricate relationship between osteoporosis and nutrition. Furthermore, the high prevalence of osteoporosis in the population and the associated societal implications have driven a surge in related research[6,11,29]. In light of this trend, it is imperative to enhance interdisciplinary collaboration, foster communication and cooperation among disciplines such as nutrition, orthopedics, and endocrinology, and facilitate the comprehensive development of relevant research. Additionally, it is advisable to encourage large-scale, long-term tracking studies to acquire more robust data and definitive conclusions. Last, it is crucial to strengthen the popularization of scientific knowledge and raise public awareness regarding the intricate interplay between osteoporosis and nutrition.

The scientific research variations and distribution of osteoporosis and nutrition in different countries are more comprehensively depicted in the country’s regional distribution map. The highest number of published papers is attributed to the United States, followed by China and South Korea, collectively contributing to two-thirds of the total articles. This observation emphasizes the United States’ research prowess and influence, establishing them as the leading force in the field of osteoporosis and nutrition. Our results illustrate that relevant articles emerged earlier in the United States than in other countries, with a relatively stable publication rate. Our research findings further highlight extensive collaborative exchanges between the United States and numerous countries. On the other hand, in recent years, China, South Korea, the United Kingdom, Canada, and other countries have witnessed an increase in article publications. This trend signifies a heightened frequency and proximity of academic exchanges between countries and regions, indicating persistent attention toward osteoporosis and nutrition research and an ongoing positive developmental trajectory.

The preeminent position in published papers is held by Seoul National University in South Korea, closely followed by Harvard University and the University of California, underscoring their exceptional contributions in this field. These teams. Remarkably skilled professors and research teams specializing in related disciplines such as nutrition and orthopedics lend support to large-scale research projects and facilitate the attainment of high-level academic accomplishments. Additionally, these universities may enjoy robust collaborative ties with internationally renowned medical institutions, research organizations, and industry partners, augmenting research cooperation opportunities and resource backing. The majority of the top 10 institutions originate from the United States, followed by South Korea and Canada. The institutional cooperation chart vividly illustrates the establishment of dynamic collaboration networks among these research institutions, fostering ample avenues for academic exchange and learning. To propel the comprehensive advancement of this field, domestic research institutions and other nations should fortify their cooperation and exchanges with these institutions.

Osteoporosis International holds the top position in terms of citation volume, exemplifying its profound academic advantage and influence in the field of osteoporosis and nutrition. The second- and third-highest-ranked journals, namely, the Journal of Bone and Mineral Research and the American Journal of Clinical Nutrition, are significantly different in the realms of nutrition and orthopedics, implying a close interconnection between osteoporosis and nutrition. Moreover, The Lancet and The New England Journal of Medicine, both of which rank among the top 10 cited journals in this field, are renowned members of the four major medical journals and have immense influence in the medical domain. This clear evidence underscores the criticality of research on osteoporosis and nutrition in advancing human society.

The research field’s individual scholars and their collaborations are clearly displayed through the author’s collaborative mapping, providing a more visual representation for prolific authors. Additionally, the academic influence of cited authors can be evaluated based on their frequency of citation, thereby reflecting their academic ability and recognition among scholars [30]. Investigating authors with high publication productivity and citation rates is particularly helpful in identifying emerging trends in this field. For instance, the scholar Looker, A C, has published numerous papers in this field that have been cited multiple times. Our findings indicate that authors with high publication volumes are predominantly located in China, whereas those with high citation frequencies are mainly affiliated with research institutions in the United States and the United Kingdom. This can be partially attributed to the earlier initiation of relevant research by Western countries, leading to the higher authority of their respective institutions. As a result, China’s citation rates have been comparatively lower due to its relatively recent involvement in the field. Nevertheless, these findings also suggest that Chinese scholars have made continuous progress in advancing research in this area in recent years. They have been actively strengthening both domestic and international collaboration, which will undoubtedly contribute to their future development in a positive manner.

Citation frequency plays a critical role in assessing the influence and quality of papers, serving as an indicator to evaluate the influence status and scientific research quality across different countries, institutions, or individuals[31]. Highly cited research signifies a valuable source of scientific knowledge within any research field. It not only reflects the research level and development trends but also serves as a scientific foundation for exploring research frontiers and hotspots[18]. An exemplary article in terms of citation frequency is “Osteoporosis”[19], which provides a comprehensive exploration of fracture risk assessment and widens the scope of treatment options to prevent fractures. This study introduces a fracture risk algorithm that integrates clinical risk factors with bone density and is effectively utilized in clinical practice for treating high-risk populations. Furthermore, the identification of crucial pathways regulating bone resorption and formation has unveiled new therapeutic strategies with distinct mechanisms of action. Osteoporosis is a chronic disease that requires long-term treatment, sometimes even lifelong treatment. It is proposed that the high-risk population for fractures cannot receive sufficient treatment, and strategies to address this treatment gap, such as widely implementing fracture liaison services and improving treatment compliance, are important challenges for the future. The article also explored the controversial relationship between nutritional factors such as calcium, vitamin D, protein, and osteoporosis[32–35]. Therefore, osteoporosis remains a serious public health issue that requires in-depth exploration and resolution by researchers. In addition, the highly cited articles “European guidance for the diagnosis and management of osteoporosis in postmenopausal women”[21] and “Vitamin D deficiency”[22] deserve the attention of researchers. These two articles discuss the close relationship between osteoporosis and postmenopausal women, as well as vitamin D. Kanis, J.A. and Holick, M.F., whose publications and cited frequencies also take the lead. Therefore, they have a profound impact on this field.

Keyword co-occurrence refers to the presence of different keywords within the same literature[36]. A high frequency of keyword occurrences indicates research hotspots and developmental trends in the field[37]. Our findings demonstrate that studies focusing on osteoporosis and nutrition prioritize bone density, women (including postmenopausal women), vitamin D, body mass index, risk, and hip fractures. Osteoporosis is distinguished by a reduction in bone mass and the deterioration of bone tissue microarchitecture, which increases the susceptibility to bone fragility and fractures[38]. In patients without fragility fractures, low bone mineral density (BMD) is often utilized to diagnose osteoporosis. However, measuring BMD via DXA is an imperfect predictor of fracture risk, as it identifies less than half of individuals who subsequently experience an osteoporotic fracture. Therefore, further exploration is needed in future research to determine whether there are better methods for measuring bone density and predictive indicators for fracture risk. A decrease in estrogen appears to be a prominent mechanism in the development of osteoporosis, particularly during menopause[39]. These findings align with our own research. However, it is important to note that there is currently limited research available specifically regarding osteoporosis in males. Insufficient evidence exists regarding the effectiveness of therapies aimed at preventing fractures or treating osteoporosis in men, primarily due to the scarcity of relevant published studies. In addition, the investigation of “risk factors” encompassing a range of characteristics. These factors include but are not limited to, advancing age, female gender, postmenopausal status, reduced gonadal function or premature ovarian failure, low body weight, a familial history of hip fractures, racial background (with a higher risk observed among individuals of white ethnicity compared to black ethnicity), prior occurrences of clinical or morphometric spinal fractures, previous fractures resulting from minor trauma (referred to as osteoporotic fractures), rheumatoid arthritis, current tobacco use, alcohol consumption (at least 3 cups per day), low bone density (BMD), inadequate vitamin D levels, insufficient calcium intake, overweight, susceptibility to falls, and fixation procedures[40]. These risk factors all include the keyword hotspots we are exploring, so they are still worthy of attention in future research. One study showed that calcium plus vitamin D3 reduced the risk for fracture among elderly women but not elderly men[41]. There is still controversy over the evidence that vitamin D and calcium supplementation reduce the risk of osteoporosis. While osteoporosis can affect any bone, certain sites, such as the hip, spine, and wrist, are especially susceptible to osteoporosis[38]. Fractures represent a significant public health concern due to their association with morbidity, functional impairment, reduced quality of life, and even mortality[8]. Consequently, comprehensive and extensive research to address the numerous problems associated with osteoporosis is crucial.

Keyword clustering has facilitated the identification of emerging research areas in this field, encompassing physical activity, machine learning, sarcopenia, trabecular bone, nutrition examination, and quality of life. Physical activity, also referred to as exercise, plays a pivotal role in promoting healthy aging by preventing or alleviating falls, pain, sarcopenia, osteoporosis, and cognitive impairment[42]. Osteoporosis and sarcopenia are diseases that commonly affect older adults and impact the musculoskeletal system. These conditions are characterized by reduced bone density and muscle mass and strength, ultimately diminishing both mobility and quality of life[39]. Numerous publications[43–47] have demonstrated the preventive benefits of resistance and endurance exercises against osteoporosis and sarcopenia. In postmenopausal women, long-term resistance or aerobic exercise is known to contribute to increased bone formation and mass[48,49]. Within the realm of biomedical research, machine learning is an advanced scientific methodology that is widely employed. For instance, a recent study conducted by Liu et al. (2022)[50] employed bioinformatics and machine learning methods to investigate crosstalk genes between periodontitis (PD) and osteoporosis (OP) and elucidated potential associations between crosstalk genes and pyroptosis-related genes. These discoveries pave the way for further investigations in the field. In addition, nutritional examination and quality of life are significant considerations in various studies. It has been established that the loss of muscle mass observed in sarcopenia contributes to an increase in insulin resistance, thereby promoting the development of metabolic syndrome and obesity[51]. Consequently, the quality of life of affected individuals declines. The consumption of milk and dairy products has been associated with a reduced risk of osteoporosis. Additionally, protein and vitamin D supplementation analysis[1] concluded that dairy consumption is not associated with the prevention of osteoporosis or fractures. Furthermore, randomized controlled trials (RCTs) have failed to confirm a decrease in the incidence of falls and fractures following vitamin D supplementation[52]. Therefore, there remains controversy regarding the relationships among various nutrients, nutritional supplements, and osteoporosis, necessitating further exploration. Finally, a keyword citation explosion analysis was performed, yielding results in line with keyword co-occurrence and clustering. The current research landscape continues to emphasize postmenopausal women, as well as topics such as vitamin D, calcium intake (including dietary calcium), and biochemical markers. Exploring the effects of exercise and nutrition on osteopenia offers potential avenues for mitigating the biomarkers involved in these two syndromes, which collectively contribute to fractures and muscle degeneration[39].

### Limitations

This study has several limitations that warrant acknowledgment. First, the bibliometric analysis focused exclusively on publications extracted from the WOSCC database, potentially overlooking influential documents. Hence, future research could enhance the comprehensiveness of the analysis by supplementing it with additional databases. Second, the retrieved data may include manuscripts currently under evaluation. Third, to present the bibliometric characteristics of the original articles, we intentionally excluded reviews from the study that inadvertently obscured some emerging trends in the field. Additionally, we confined our search to articles published between January 2004 and February 2024, excluding those published outside this timeframe. Lastly, the pool of recruited manuscripts may feature weaker or peripheral works, potentially distorting the analysis to some extent.

### Conclusions

Using the WOSCC database, we used CiteSpace to analyze osteoporosis and nutrition-related articles published from 2004 to 2024, aiming to identify publication patterns, contributors, and recent research trends. The analysis revealed a consistent annual increase in productivity, particularly since 2018, indicating a predicted continuation of this upward trend. To effectively respond to this trend, it is imperative to prioritize strengthening interdisciplinary collaboration, fostering communication and cooperation among disciplines such as nutrition, orthopedics, and endocrinology, and facilitating comprehensive research development. The examination of publication origins indicated that the United States was the leading contributor, with Seoul National University (SNU) emerging as the most productive institution. Extensive collaborative exchanges among these countries and institutions highlight persistent attention and positive growth in osteoporosis and nutrition research. Authors with high publication volumes are predominantly based in China, while those with significant research in this field occurred relatively late, but Chinese scholars have actively pursued domestic and international collaborations, which undoubtedly augment their future development in a constructive manner. Research hotspots include areas such as bone density, postmenopausal women, vitamin D, hip fractures, etc. Research subjects encompass various aspects, such as physical activity, sarcopenia, calcium intake, body mass index, etc. Notably, current research still has some limitations that warrant further attention and exploration. Future studies should prioritize the exploration of improved methods to measure bone density and predictive indicators of fracture risk. Additionally, there is a paucity of research specifically focusing on osteoporosis in males. Controversies persist regarding the relationships among various nutrients, nutritional supplements, and osteoporosis, necessitating further investigation. The findings from such research endeavors will aid investigators in assessing the current landscape and identifying novel avenues for future exploration within this field. Finally, as osteoporosis poses a significant clinical challenge to the global population, undertaking comprehensive and extensive research is crucial to address the numerous complexities associated with this condition.

## Data Availability

All relevant data are within the manuscript and its Supporting information files.

## Aknowledgments

We would like to appreciate those who provided help in data collecting and paper writing.

## Author Contributions

**Conceptualization:** Min Li and Binyang Yu

**Data curation:** Haiyan Yang and Haiyan He

**Formal analysis:** Rui Gao, Ning Li, Aili Lv and Xiaoling Zhou

**Funding acquisition:** Rui Gao

**Investigation:** Haiyan Yang and Haiyan He

**Methodology:** Min Li and Binyang Yu

**Project administration:** Rui Gao, Ning Li, Aili Lv and Xiaoling Zhou

**Resources:** Min Li and Binyang Yu

**Software:** Min Li and Binyang Yu

**Supervision:** Rui Gao, Ning Li, Aili Lv and Xiaoling Zhou

**Visualization:** Min Li and Binyang Yu

**Writing—original draft preparation:** Min Li, Binyang Yu, Haiyan Yang and Haiyan He

**Writing—review and editing:** Min Li, Binyang Yu and Rui Gao

